# The Impact of State Paid Sick Leave Policies on Longitudinal Weekday Workplace Mobility During the COVID-19 Pandemic

**DOI:** 10.1101/2022.01.16.22269377

**Authors:** Catherine C. Pollack, Akshay Deverakonda, Fahim Hassan, Syed Haque, Angel N. Desai, Maimuna S. Majumder

**Affiliations:** COVID-19 Dispersed Volunteer Research Network; Boston, Massachusetts, USA; Department of Biomedical Data Science, Geisel School of Medicine at Dartmouth; Lebanon, New Hampshire, USA; Department of Epidemiology, Geisel School of Medicine at Dartmouth; Lebanon, New Hampshire, USA; School of Public Health, University of Alberta; Edmonton, Alberta, Canada; Network Science Institute, Northeastern University; Boston, Massachusetts, USA; Department of Internal Medicine, Division of Infectious Diseases, University of California - Davis; Sacramento, California, USA; Computational Health Informatics Program, Boston Children’s Hospital; Boston, Massachusetts, USA; Department of Pediatrics, Harvard Medical School; Boston, Massachusetts, USA

**Keywords:** COVID-19, Paid Sick Leave, Physical Distancing, Workplace Mobility, Health Policy

## Abstract

**Objectives:** To evaluate whether the Families First Coronavirus Response Act (FFCRA) modified the association between pre-existing state paid sick leave (PSL) and weekday workplace mobility between February 15 and July 7, 2020.

**Methods:** The 50 US states and Washington, D.C. were divided into exposure groups based on the presence or absence of pre-existing state PSL policies. Derived from Google COVID-19 Community Mobility Reports, the outcome was measured as the daily percent change in weekday workplace mobility. Mixed-effects, interrupted time series regression was performed to evaluate weekday workplace mobility after the implementation of the FFCRA on April 1^st^, 2020.

**Results:** States with pre-existing PSL policies exhibited a greater drop in mobility following the passage of the FFCRA (**β**=-8.86,95%CI:-11.6,-6.10,*P*< 001). This remained significant after adjusting for state-level health, economic, and sociodemographic indicators (**β**=-3.13,95%CI:-5.92,-0.34,*P*=.039).

**Conclusions:** Pre-existing PSL policies contributed to a significant decline in weekday workplace mobility after the FFCRA, which may have influenced local health outcomes.

**Policy implications:** The presence of pre-existing state policies may differentially influence the impact of federal legislation enacted during emergencies.

## INTRODUCTION

The COVID-19 pandemic has upended every facet of life, necessitating systemic policies to reduce its spread. Despite the deployment of COVID-19 vaccines, the ability to quarantine and isolate after exposure remains critical in order to minimize both the potential for “breakthrough cases” and the risk of infection for those who are unvaccinated. ^1^ One policy to facilitate self-quarantine and self-isolation is paid sick leave (PSL), which allows employees to take compensated time off from work to recover from illness or injury. PSL has previously been associated with a three-fold increase in protection of workers’ jobs, income, and health while recovering from illness. ^2^ PSL is especially crucial during outbreaks of communicable diseases as it can help mitigate “presenteeism,” whereby employees go to work even if they are sick.^3^ This is particularly important for COVID-19 since individuals can present a range of symptoms, including being asymptomatic or having mild symptoms akin to other, less severe respiratory illnesses.

The United States (U.S.) is one of only two Organisation for Economic Co-operation and Development countries that does not have a nationwide PSL policy, resulting in a patchwork system that varies between states. ^2,4^ Within each state, access to PSL is associated with many factors, including industry type, race, ethnicity, gender, sexual orientation, income level, immigration status, company size, full-or-part time status, and experience level. As a result, up to 40% of American private sector workers, including 69% of the lowest quartile of wage earners, are not afforded PSL. ^5^ This was partially rectified with the Families First Coronavirus Response (FFCRA) and Coronavirus Aid, Relief and Economic Security (CARES) Acts, which provided emergency, two-week PSL on April 1st, 2020. ^6^ This federally-legislated PSL has played an important role in slowing the spread of COVID-19 in the workplace by allowing for self-quarantine and self-isolation from work environments. ^6,7^ However, exemptions for certain employee categories (e.g., health care workers and emergency responders) and businesses with more than 500 employees blunted its coverage to potentially as few as 47% of private-sector workers. ^7^ Thus, the presence of pre-existing state PSL may have influenced how this emergency federal legislation impacted key outcomes such as travel to-and-from the workplace (i.e., weekday workplace mobility), which could be considered a proxy for workplace presenteeism and absenteeism. ^8^ As a result, it is critical to identify the differential impacts of the FFCRA on states that had pre-existing state PSL in order to elucidate what fundamental level of local preparedness is required to maximize the impact of federal legislation. The purpose of this study was to explore the impact of pre-existing state PSL on weekday workplace mobility during the time period surrounding the passage of the FFCRA (i.e., February through July 2020). It was hypothesized that states that had pre-existing state PSL would experience a greater drop in weekday workplace mobility compared to states that did not.

## MATERIALS AND METHODS

### Data collection

Four data sets were integrated for each of the 50 states and Washington, DC. The primary exposure of interest (i.e., presence or absence of pre-existing state PSL) was coded as either “yes” or “no” based on data from the Kaiser Family Foundation. ^4^ The primary outcome of interest (i.e., weekday workplace mobility) was collected from Google COVID-19 Community Mobility Reports. ^9^ Within these reports, weekday workplace mobility was calculated as the percent change in mobility between the date of interest and a pre-pandemic baseline. This baseline was computed as the median mobility between January 3 and February 6, 2020 on the same day of the week (e.g., Monday, Tuesday) as the date of interest. Economic covariates (e.g., wage policies, worker protection policies, right-to-organize policies) and epidemiological metrics (e.g., COVID-19 cases and deaths per state) were collated from the Oxfam Index and the *New York Times* COVID-19 database, respectively. Other sociodemographic factors (e.g., median household income, state gross domestic product [GDP], commuting patterns, presidential election results between 2004 and 2016) were gathered from the American Community Survey and the Federal Election Commission. ^10–13^

### Statistical analysis

A mixed-effects, interrupted time series regression model with nested random effects for state and month characterized the relationship between the presence of pre-existing state PSL and daily percent change in weekday workplace mobility. The initial model only adjusted for temporality relative to the implementation of the FFCRA on April 1st, 2020 (i.e., days pre-FFCRA, instantaneous FFCRA, and days post-FFCRA). Additional bivariate analyses were performed to identify which covariates were significantly associated with weekday workplace mobility. Highly correlated terms were evaluated by investigators to determine which should be retained for further analysis. A multivariable model was subsequently constructed using the same underlying structure as the unadjusted model and included all terms that were significant in the bivariate analysis. Data were aggregated with Python (version 3.8) and analyzed in R (version 4.0.3) using the RStudio Integrated Development Environment (version 1.3.1093).

## RESULTS

Immediately after the implementation of the FFCRA on April 1st, 2020, Washington DC and the 12 states with pre-existing state PSL experienced an 8.86 percentage point greater decrease in weekday workplace mobility (**β** = -8.86, 95% CI: -11.6, -6.10, *P* < .001) compared to the 39 states that do not have pre-existing state PSL (Fig. 1). Health indicators associated with a greater decrease in mobility included new cases per 100,000 (**β** = -0.03, 95% CI: -0.04, -0.03, *P* < .001) and new deaths per 100,000 (**β** = -0.43, 95% CI: -0.51, -0.35, *P* < .001). The majority of travel metrics were associated with weekday workplace mobility, although directionality varied. For example, while average commute time was inversely associated with weekday workplace mobility (**β** per minute = -1.04, 95% CI: -1.22, -0.86, *P* < .001), the percent commuting via carpool was associated with an increase in weekday workplace mobility (**β** = 1.73, 95% CI: 0.63, 2.83, *P* = .003). The bulk of economic indicators were also associated with weekday workplace mobility, including 2017 median household income (**β** per $10,000 USD = -2.47, 95% CI: -3.64, -1.29, *P* < .001) and unemployment rate (**β** = -0.31, 95% CI: -0.40, -0.20, *P* < .001). In addition, states with a dominant labor sector in “education and health services” had a greater drop in weekday workplace mobility compared to states with a dominant labor sector in “trade, transportation, and utilities” (**β** = -4.90, 95% CI: -9.39, -0.42, *P* = .044). Several demographic indicators were also associated with weekday workplace mobility, albeit in various directions. For example, while a higher percentage of men was associated with an increase in weekday workplace mobility (**β** = 2.83, 95% CI: 1.11, 4.55, *P* = .002), a higher percentage of Asian individuals was associated with a greater decrease in weekday workplace mobility (**β** = -0.31, 95% CI: -0.58, -0.05, *P* = .024). In terms of policies, states that provided paid *family* leave had a greater drop in weekday workplace mobility compared to states that did not (**β** = -10.6, 95% CI: -14.8, -7.02, *P* < .001). Finally, a higher state population per square mile was associated with a greater drop in weekday workplace mobility (**β** per 1,000 persons = -2.04, 95% CI: -2.84, -1.23, *P* < .001). See Supplementary Table 1 for a comprehensive list of covariates.

**Fig 1.**
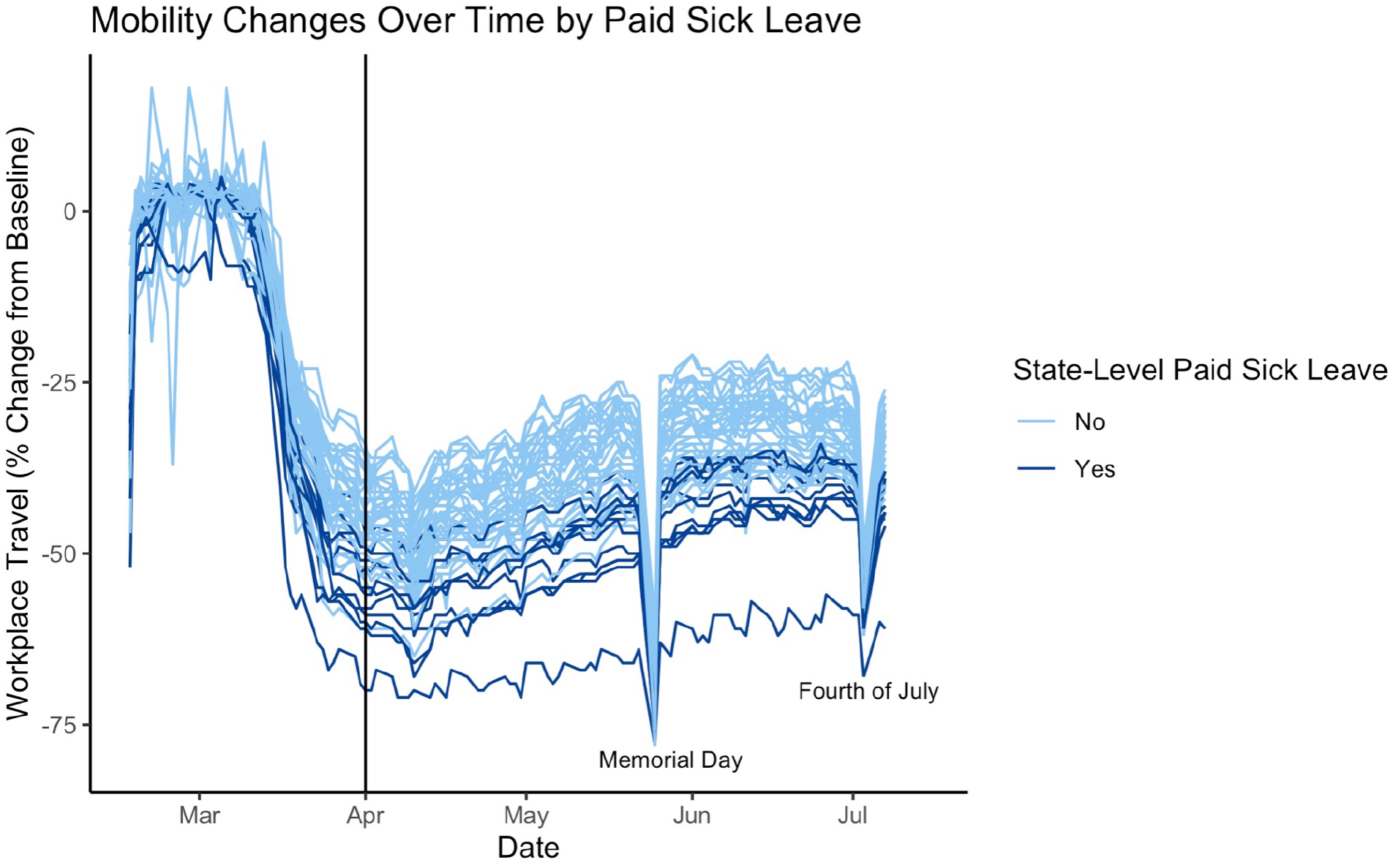
Changes in workplace travel over time by state-level paid sick leave. The black line on April 1, 2020 denotes the implementation of the Families First Coronavirus Response Act (FFCRA). Twelve states (Arizona, California, Connecticut, the District of Columbia, Massachusetts, Maryland, New Jersey, New York, Oregon, Rhode Island, Vermont, and Washington) had pre-existing paid sick leave policies mandated by the state, whereas the remaining 39 did not. The most substantial drops occurred on two federal U.S. holidays: Memorial Day (May 25th, 2020) and Independence Day (July 4th, 2020)

After adjusting for these and other covariates, the association between pre-existing state PSL and weekday workplace mobility remained statistically significant (**β**_paid sick leave_ = -3.13, 95% CI: -5.92, -0.34, *P* = .039; Table 1). Other variables that retained their significance and were associated with a decrease in weekday workplace mobility included new cases per 100,000 (**β** = -0.03, 95% CI: -0.04, -0.03, *P* < .001), average commute time (**β** per minute = -0.59, 95% CI: -0.94, -0.24, *P* = .004), unemployment rate (**β** = -0.35, 95% CI: -0.45, -0.26 *P* < .001), and state population per square mile (**β** per 1,000 persons = -1.12, 95% CI: -2.04, -0.20, *P* = .027). Variables that retained their significance and were associated with an increase in weekday workplace mobility included poverty rate (**β** = 0.50, 95% CI: 0.07, 0.94, *P* = .035) and having “manufacturing” as a dominator labor sector relative to “trade, transportation, and utilities” (**β** = 7.34, 95% CI: 0.59, 14.1, *P* = .045). See Table 1 for the full model.

**Table 1.**
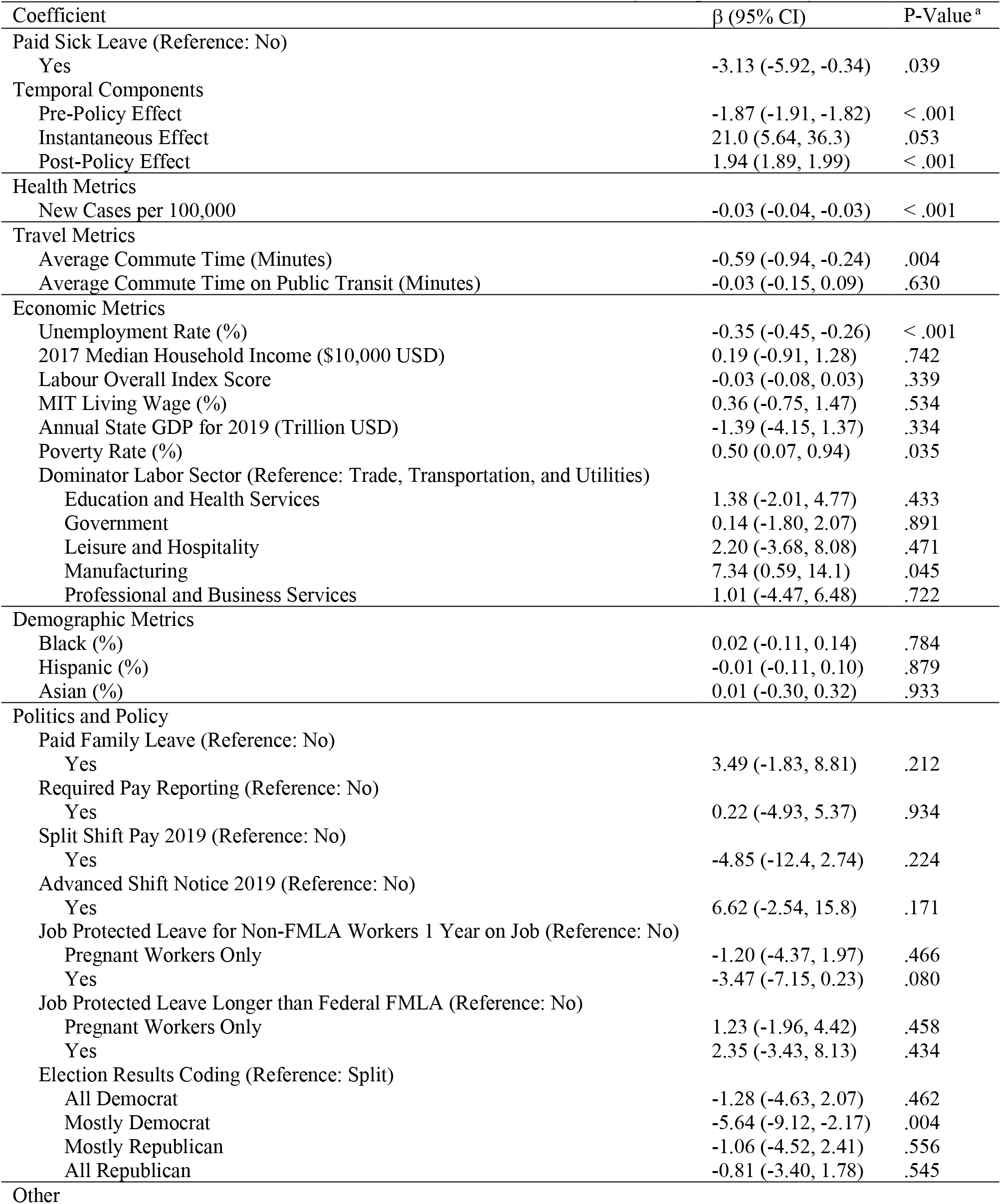
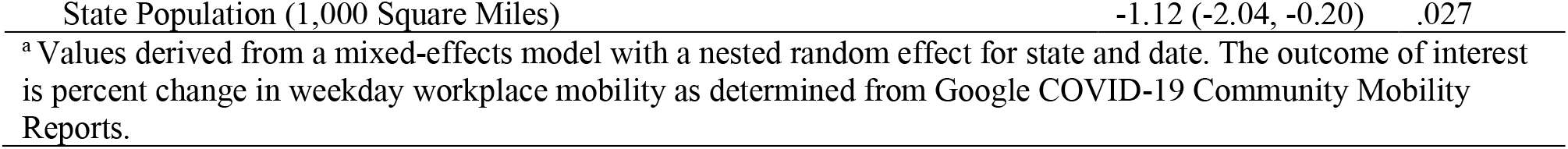
Multivariable Mixed Effects Model: Paid Sick Leave vs. Weekday Workplace Mobility

## DISCUSSION AND CONCLUSIONS

This study is the first to comprehensively evaluate the impact of pre-existing state PSL on weekday workplace mobility in the U.S. during the COVID-19 pandemic. The presence of pre-existing state PSL was significantly associated with a drop in weekday workplace mobility in the early phase of the pandemic in both unadjusted and adjusted models. The results presented here suggest a complex interplay between pre-existing labor workforce protections and emergency public health interventions targeted for the workforce.

Increasingly, states are considered the primary vehicles for managing and administering social services, leading to highly variable policies. ^14^ Thus, the presence of pre-existing state PSL acted as a “classifier” that could differentiate how the FFCRA impacted weekday workplace mobility at the state level. Given the ubiquity of COVID-19, this evaluation provides a nationwide, ecological assessment that may suggest that federal emergency aid packages have a stronger impact in states and other localities with the pre-existing infrastructure to support such policies. More specifically, this study also contributes to the growing literature characterizing the impact of the FFCRA and its emergency PSL on various health and behavioral outcomes. A prior study, which relied on cellular data in place of Google COVID-19 Community Mobility Reports, also found that the FFCRA significantly decreased the time spent away from home. However, the FFCRA’s impact on *workplace* mobility––as is the focus of this study–– could not be determined. ^8^

As COVID-19 variants of concern continue to emerge, the lack of consistency in PSL policies across the U.S. leaves employees vulnerable, especially those who are considered “essential workers” or are in positions that require in-person work. ^15^ This disproportionately impacts Black, Indigenous, People of Color as well as the socioeconomically disadvantaged–– the same groups that are both at higher risk for COVID-19 and disenfranchised by current labor laws. ^16^ To protect such individuals, there is a need for permanent structural changes in labor protection laws at the federal level, which could leverage pre-existing state policies to identify best practices and potential pitfalls. ^17^ Furthermore, systematic changes to labor protection laws could also contribute in the long term to improving preparedness in emergency situations as well as overall social and health equity.

As a social determinant of health, the presence or absence of PSL has ramifications for one’s health, well-being, and quality of life ^18,19^. Having PSL makes an employee 60% more likely to receive an influenza vaccination and engage with medical and cancer screenings without needing to consider forfeiting their income or jobs. ^3^ Moreover, an additional study found that people without PSL were three times as likely to delay/not seek at all needed treatment, due to likely concerns about the immediate costs of said treatment and related costs of wage loss. ^20^ PSL’s unique role as a social determinant of health is further underscored by this trend holding constant when controlling for health status, education level, and income level. ^20^ The impact of PSL’s presence or absence also applies to immediate family members as well; parents who had PSL were more likely to take time off to care for children when needed. ^21^ Low-income children were less likely to have parents who had PSL, and even within this group there were further differences; lowest-income children were even less likely to have a parent with PSL, and similarly with low-income children with health problems.^21^ The effects of this social determinant for an individual also extend to the community at large; one study estimated that due to a lack of PSL, 7 million people were additionally infected by people exhibiting “presenteeism” in the workplace during the H1N1 pandemic. ^22^ On the other hand, one study estimated that Connecticut’s PSL law resulted in a 14.8% reduction in the spread of illness in 2013. ^23^

While this study is the first to examine the impact of pre-existing state PSL on weekday workplace mobility during the COVID-19 pandemic, it is not without its limitations. First, publicly available covariate data were compiled across multiple sources and may have been measured at different points in time; thus, future work should attempt to standardize the time frame of analysis. Second, analysis was limited to the early stages of the COVID-19 pandemic, presenting opportunities for examination of the long-term impacts of pre-existing state PSL on workplace mobility and other metrics. Third, given the ecological nature of the study, future work is necessary to quantify the direct, person-level impact of pre-existing state PSL on adherence to workplace mobility measures. Fourth, Google COVID-19 Community Mobility Reports may not be representative of all populations (e.g., those without access to a cellular device), and the calculation of daily changes relative to a baseline in January and February 2020 (as opposed to a full year) may result in some seasonal biases. This may bias results away from the null, as individuals may be less likely to take off work during January and February compared to the following months. Finally, this study is limited to PSL, and evaluation of additional economic policies––such as medical leave for family members, flexible work hours, remote work policies, and flexibility in shift work––could offer more nuanced perspectives.

PSL is fundamental to preserving the health of the workforce, particularly during times of crisis. The results presented here suggest that pre-existing state policies may enhance the effectiveness of emergency legislation, although long-term, systemic labor protection laws remain crucial. Successful implementation of such laws requires an equity-based approach that considers addressing disparities in access to labor benefits, thoughtful outreach strategies through clear and consistent communication to all labor force members, and rigorous oversight and enforcement from state and federal labor departments and boards to both ensure compliance by employers and maximize the potential for success. ^17^

## Supporting information

STROBE Checklist

ICMJE Conflict of Interest Form

## Data Availability

All data produced in the present study are available upon request.

