## Supplementary Table 1 for "The Impact of State Paid Sick Leave Policies on Longitudinal Weekday Workplace Mobility During the COVID-19 Pandemic"

### Supplementary Information

Supplementary Table 1. Bivariate covariate analysis
Note that each row represents an independent model.

| Bivariate Comparisons Model | | |
|---|---|---|
| Coefficient | β (95% CI) | P-Value [a] |
| Health Metrics | | |
| Cumulative Cases per 100 | -5.41 (-6.36, -4.45) | < .001 |
| New Cases per 100,000 | -0.03 (-0.04, -0.03) | < .001 |
| New Deaths per 100,000 | -0.43 (-0.51, -0.35) | < .001 |
| Travel Metrics | | |
| Travel to Transit Stations (% Change from Baseline) | 0.42 (0.40, 0.44) | < .001 |
| Average Commute Time (Minutes) | -1.04 (-1.22, -0.86) | < .001 |
| Average Commute Time on Public Transit (Minutes) | -0.39 (-0.58, -0.19) | < .001 |
| Commute via Car/Truck/Van Alone (%) | 0.49 (0.35, 0.62) | < .001 |
| Commute via Car/Truck/Van Carpool (%) | 1.73 (0.63, 2.83) | .003 |
| Commute via Public Transportation Excluding Taxi (%) | -0.69 (-0.86, -0.53) | < .001 |
| Commute via Walking (%) | -1.35 (-2.09, -0.62) | < .001 |
| Commute via Other Means (%) | -2.41 (-3.95, -0.87) | .003 |
| Work at Home (%) | -1.40 (-2.83, 0.02) | .057 |
| Economic Metrics | | |
| Unemployment Rate (%) | -0.31 (-0.40, -0.20) | < .001 |
| 2017 Median Household Income ($10,000 USD) | -2.47 (-3.64, -1.29) | < .001 |
| Labour Overall Index Score | -0.15 (-0.20, -0.10) | < .001 |
| MIT Living Wage (%) | -1.87 (-2.42, -1.31) | < .001 |
| Annual State GDP for 2019 (Trillion USD) | -3.65 (-6.40, -0.99) | .012 |
| Poverty Rate (%) | 0.65 (0.08, 1.22) | .028 |
| Dominator Labor Sector (Reference: Trade, Transportation, and Utilities) | | |
| Education and Health Services | -4.90 (-9.39, -0.42) | .044 |
| Government | -2.13 (-6.19, 1.93) | .326 |
| Leisure and Hospitality | -3.55 (-11.4, 4.28) | .395 |
| Manufacturing | 0.90 (-9.99, 11.8) | .876 |
| Professional and Business Services | -4.23 (-15.1, 6.67) | .466 |
| Demographic Metrics | | |
| Male (%) | 2.83 (1.11, 4.55) | .002 |
| Black (%) | -0.13 (-0.28, 0.01) | .081 |
| Hispanic (%) | -0.17 (-0.32, -0.02) | .026 |
| Asian (%) | -0.31 (-0.58, -0.05) | .024 |
| 65 and Above (%) | 0.59 (-0.18, 1.37) | .137 |
| Politics and Policy | | |
| Paid Family Leave (Reference: No) | | |
| Yes | -10.6 (-14.8, -7.02) | < .001 |
| Required Pay Reporting (Reference: No) | | |
| Yes | -8.48 (-11.9, -5.07) | < .001 |
| Split Shift Pay 2019 (Reference: No) | | |
| Yes | -9.55 (-14.2, -4.94) | < .001 |
| Advanced Shift Notice 2019 (Reference: No) | | |
| Yes | -9.72 (-15.0, -4.49) | < .001 |
| Job Protected Leave for Non-FMLA Workers 1 Year on Job (Reference: No) | | |
| Pregnant Workers Only | -1.23 (-5.58, 3.11) | .581 |

| | | |
|---|---|---|
| Yes | -6.84 (-10.1, -3.54) | < .001 |
| Job Protected Leave Longer than Federal FMLA (Reference: No) | | |
| Pregnant Workers Only | -1.88 (-7.37, 3.63) | .510 |
| Yes | -7.77 (-13.3, -2.27) | .008 |
| Election Results Coding (Reference: Split) | | |
| All Democrat | -6.63 (-11.5, -1.73) | .012 |
| Mostly Democrat | -4.52 (-9.93, 0.90) | .117 |
| Mostly Republican | 0.07 (-7.09, 7.24) | .985 |
| All Republican | 2.32 (-2.53, 7.16) | .364 |
| Flexible Scheduling (Reference: No) | | |
| Yes | -2.43 (-9.15, 4.29) | .478 |
| Other | | |
| State Population (1,000 Square Miles) | -2.04 (-2.85, -1.23) | < .001 |
| US State Land Area Square Miles (Million Square Miles) | 16.3 (-1.88, 34.5) | .083 |

[a] Values derived from a mixed-effects model with a nested random effect for state and date. The outcome of interest is percent change in workplace mobility as determined from Google COVID-19 Community Mobility Reports
